# Susceptibility and Sustainability of India against CoVid19: a multivariate approach

**DOI:** 10.1101/2020.04.16.20066159

**Authors:** Soumi Ray, Mitu Roy

## Abstract

**Purpose:** We are currently in the middle of a global crisis. Covid19 pandemic has suddenly threatened the existence of human life. Till date, as no medicine or vaccine is discovered, the best way to fight against this pandemic is prevention. The impact of different environmental, social, economic and health parameters is unknown and under research. It is important to identify the factors which can weaken the virus, and the nations which are more vulnerable to this virus.

**Materials and Methods:** Data of weather, vaccination trends, life expectancy, lung disease, number of infected people in the pre-lockdown and post-lockdown period of highly infected nations are collected. These are extracted from authentic online resources and published reports. Analysis is done to find the possible impact of each parameter on CoVid19.

**Results:** CoVid19 has no linear correlation with any of the selected parameters, though few parameters have depicted non-linear relationship in the graphs. Further investigations have shown better result for some parameters. A combination of the parameters results in a better correlation with infection rate.

**Conclusions:** Though depending on the study outcome, the impact of CoVid19 in India can be predicted, the required lockdown period cannot be calculated due to data limitation.

## Introduction

The entire world has almost stopped theoretically in the month of March 2020. This is one of the most unexpected and unbelievable situation in world’s history. A virus, starting its journey from Wuhan city of China in December 2019, has now reached almost all major cities and has created colonies very rapidly. As per World Health Organization (WHO), the first case of CoVid19 was identified on 8th December 2019 [1]. Initially the disease was misunderstood as some variation of influenza. Scientists, researchers and doctors, after doing continuous analysis, then came up with information about this novel corona virus. Though the drug is not yet in the market, the structural details of the virus are now known to us[2]. Because of its similarities with the behavior of severe acute respiratory syndrome (SARS) corona virus, this virus was named as severe acute respiratory syndrome coronavirus 2 (SARS-CoV-2) and the disease was identified as coronavirus disease (CoVid19) by WHO [3]. Seasonal diseases which have higher mortality rate usually belong to SARS category. Observing the high infection rate, on11th March 2020, WHO declared CoVid19 as pandemic [4].

In 2003, Asian countries were badly affected by SARS epidemic which originated in China. The worldwide death toll was 774, having a ratio of 1:10 of registered cases [5]. In 1957, Asian flu virus claimed around thousand lives in India[6]. One of the severest global pandemic in the history of recent past was 1918 flu. This flu was first reported in a Spanish newspaper, and it infiltrated India through Bombay port. The disease was contagious. It claimed 400 million lives worldwide [7]. It is claimed that one third of the population was infected. Different articles claimed China as the origin of the flu [8-10]. But death in China itself was very few, whereas India lost almost one fifth of its population [11]. The estimated death toll was 14 million [12]. The flu was so deadly, that it brought the population down for first time as well as the last time, till date in the history of India. But the virus disappeared almost suddenly after few months. It is assumed that like any other pathogen, this virus also rapidly mutated to a lesser lethal strain and then finally died out [5].

CoVid19 has many similarities with these 1918 flu and 2003 SARS. All are viral and contagious infections, epidemic in nature, turning into pandemic within few weeks, transmitted through droplets and very easily transmissible from human to human. It is also suspected that coronavirus can be transmitted through air and it can survive in environment without any decrease in its efficiency for a long time and thus the chance of infection increases[13]. If coronavirus is transmissible and airborne, this is an alarming situation for India.

Normally during season change, Indians suffer from different common ailments like cold and cough, nasal congestion, conjunctivitis etc. Many of these diseases are infectious and transmissible. A large part of the population suffers from one or more of these issues commonly. Along with those common epidemics, CoVid19 has to be taken care of. Due to nor’westers during March/April the chances of fast spreading of CoVid19 is also high. Many people are already infected, among which all are not having significant symptoms and hence not identified.

Finding ways to stop community transmission, case identification at initial stage and controlling the death rate is an emergency. Even the cure will not be easy to save the world if the infection is not prevented. Faster and easier worldwide transportation system has turned into a curse in case of viral epidemics. Due to international travels, almost all the countries in the world have got infected by this transmissible virus within a very short duration simultaneously. CoVid19 has affected 209 Countries and Territories around the world as of 8th April 2020.

From the information of the impact of 1918 pandemic in India, we have tried to understand the underlying facts. A large population who lived near and below poverty line got affected by the 1918 pandemic [14]. Apparently, it seems that, the sanitation had a significant relation with the disease infection, spread and severity. But the high mortality rate in case of CoVid19 even in developed countries raises a question on this easy assumption. Light from a different angle may have some answer to it.

People from lower economic segment not only failed to maintain sanitation but also suffered from improper diet. Lack of consumption of proper and healthy food weakened their immunity. Before 1918, no vaccine other than for smallpox was available. No vaccine or antibacterial was invented to prevent the pandemic diseases. It is not very difficult to assume that the practice of vaccination among the poor people in India was also low. So, hygiene was not solely responsible for the devastating death toll of 1918. Immunity also played a bigger role in it.

In this article, we have tried to give some insight from available worldwide information, in order to understand the nature of this new coronavirus infection which is causing CoVid19. We have tried to inspect its dependencies on other known parameters. A measure of the possible effects of different parameters on the outbreak and infection growth has also been estimated in order to understand the risk in India. The scientists, from different parts of world, already have done researches and have published valuable information. All the important outcomes have been considered and examined before concluding our findings. A common drawback is associated with most of the reported works. They have discussed impact of single dimensions like temperature, vaccination or life cycle of virus. This has restricted the scope of understanding of the virus’s overall activities and limited the chance of prediction and prevention. We have tried to overcome this limitation by replacing univariate analysis with multivariate approach. This article has considered different possible aspects to get a robust outcome.

## Material & methods

To keep the result as unbiased as possible, we have collected data of multiple cities and countries all over the world having different geographical locations and climatic conditions. We have conducted analysis of several relevant factors to look into the situation from all possible corners. Possible dependency of the number of total infection and death has been examined against the environmental conditions taking different weather parameters as independent variables. For this purpose, data of 43 cities all over the world has been considered. These cities are significantly affected by SARS-CoV 2. We have collected the CoVid19 related data from WHO site and other data from particular websites for each individual type of parameter to minimize the biases. The duration considered to check dependency of weather parameters is from 1st to 27^th^ March, as because by 1^st^ of March, a large number of countries got significantly infected. The days have been limited to 27^th^, as by 3^rd^ week of March majority of the countries applied social distancing and isolation. Data beyond that time may have significant impact of isolation. Isolation includes noise in the measure of infection and death, as it puts restriction in virus transmission due to lower availability of hosts.

The weather information of all those locations have been collected from a single website [15] to keep it uniform even if the information includes any noise or bias. We also have taken a measure of the population to compare the infection rate. Human to human infection depends on the population for transmission. Other than these affecting parameters, another checking has been done on the impact due to the lockdown. How the duration of lockdown has been affecting the number of new infection, have also been examined to understand its importance.

We have considered life expectancy also to inspect its impact on the number of infected cases and deaths. The life expectancy includes the impact of different parameters like average living standard, socioeconomic situation, health service qualities, natural calamities etc. A comparison with life expectancy refers to be have a relation with all those hidden parameters though the insights of each are not accessible. The data is collected from United Nations Development Program reports [16]. As proposed in a paper [17], vaccination may have great impact on death. In This article, the data of Bacillus Calmette–Guérin (BCG) vaccination has been compared with present death rate of different countries. This observation has a significant impact in our final outcome. This data is collected from WHO and review articles [18-20]. The additional factors included in this study are the impact of lung cancer (LC), chronic obstructive pulmonary disease (COPD) and Lower respiratory infect (LRI). These diseases have shown an impact on death rate in many countries which are badly affected by coronavirus. The required data are retried from online resources [21].

### Data analysis and Result

The total number of cases per million and total death per million are very much correlated as per our examination (ρ=0.69, p= 6.2E-31) as of 12^th^ April 2020. Because of the good correlation, any of these two parameters can be used for prediction analysis and the outcome will remain comparable. In this article we have used either of these parameters to understand the impact of all other parameters on Covid19 and the results are compared later.

### Impact of weather

We have divided our test result into different parts. In the first part we have discussed the impact of different weather parameters on the number of infected cases. We have not considered active cases because the number is ever changing with continuous addition of new cases, elimination of recovery numbers and deaths. Initially the number of infected cases increased rapidly due to lack of awareness, availability of more hosts, free movement of hosts etc. and then gradually decreased for enforcement of quarantine, growing awareness like washing our hands regularly, social distancing etc. These qualitative parameters are not traceable but have significant impact on the number of new cases. Different cities, selected in this article, have different geographical locations with widely varying atmospheric condition. If any of the considered weather parameters have a significant effect on infection transmission, then that will impact the transmission equally, irrespective of the location. The list of the cities with parameters details is given in the Appendix.

We have taken data per million to nullify the population bias. As the virus can be transmitted from human to human and can travel a small distance through droplets with airflow, higher population increases the availability of new hosts and hence the rate of infection. Our target is to inspect the significance of that impact. This study is very important for India as its population is very high.

### Weather impact on registered cases

The correlation between number of registered infected cases per million and the different weather parameters like temperature measures, humidity, dew points and precipitation have been found. We have used Pearson’s correlation to check the dependency of identified case numbers with the other parameters. We have used linear regression correlation to understand the linearity in the relationship, its statistical significance and to make a prediction. The results are presented in tabular form in table 1.

**Table 1:** correlation between cases per million and different weather parameters

| | Correlation($\rho$) | p-value |
| --- | --- | --- |
| Avg. of max temp. | 0.1397 | 0.3962 |
| Avg. of min. temp. | 0.1765 | 0.2825 |
| Avg. of deviation | 0.1114 | 0.4996 |
| Highest temp | 0.1198 | 0.4675 |
| Lowest temp. | 0.1697 | 0.3017 |
| Max deviation | 0.0421 | 0.7993 |
| Min deviation | 0.0873 | 0.5973 |
| Avg. max dew point | 0.1351 | 0.4122 |
| Avg. Min dew point | 0.1048 | 0.5253 |
| Avg. max humidity | 0.1145 | 0.4877 |
| Avg. min humidity | 0.0416 | 0.8016 |
| Avg. max wind speed | 0.2058 | 0.2089 |
| Avg. min wind speed | 0.1013 | 0.5394 |
| Avg. High press | 0.1544 | 0.3480 |
| Avg. low press | 0.1647 | 0.3163 |

The p-value in each case is high enough to support null hypothesis and reject any considerable correlation. But we decided to inspect further. Each parameter is examined separately to find any possible impact of CoVid19 deaths. This method offers exciting information about lowest temperature and highest temperature. We have considered death of 0.01% of population as the threshold. Above this rate is considered as a matter of concern. As per Central Intelligence Agency, present population of India is 1,326,093,247 [22]. The 0.01% of this population is about 1,32,609 which is a few thousand more than present death count all over world and not an ignorable count. The countries which experienced higher death rate (as well as death) had the minimum temperature below 0°C as shown in figure 1. The Y-axes are changed to logarithmic scale to enhance the data-point visibility. The linear trend lines are also shown in the figures for visual depiction of our understanding, that is with increase in minimum temperature the death rate and total death (hence infection rate) decreases.

**Figure 1:**
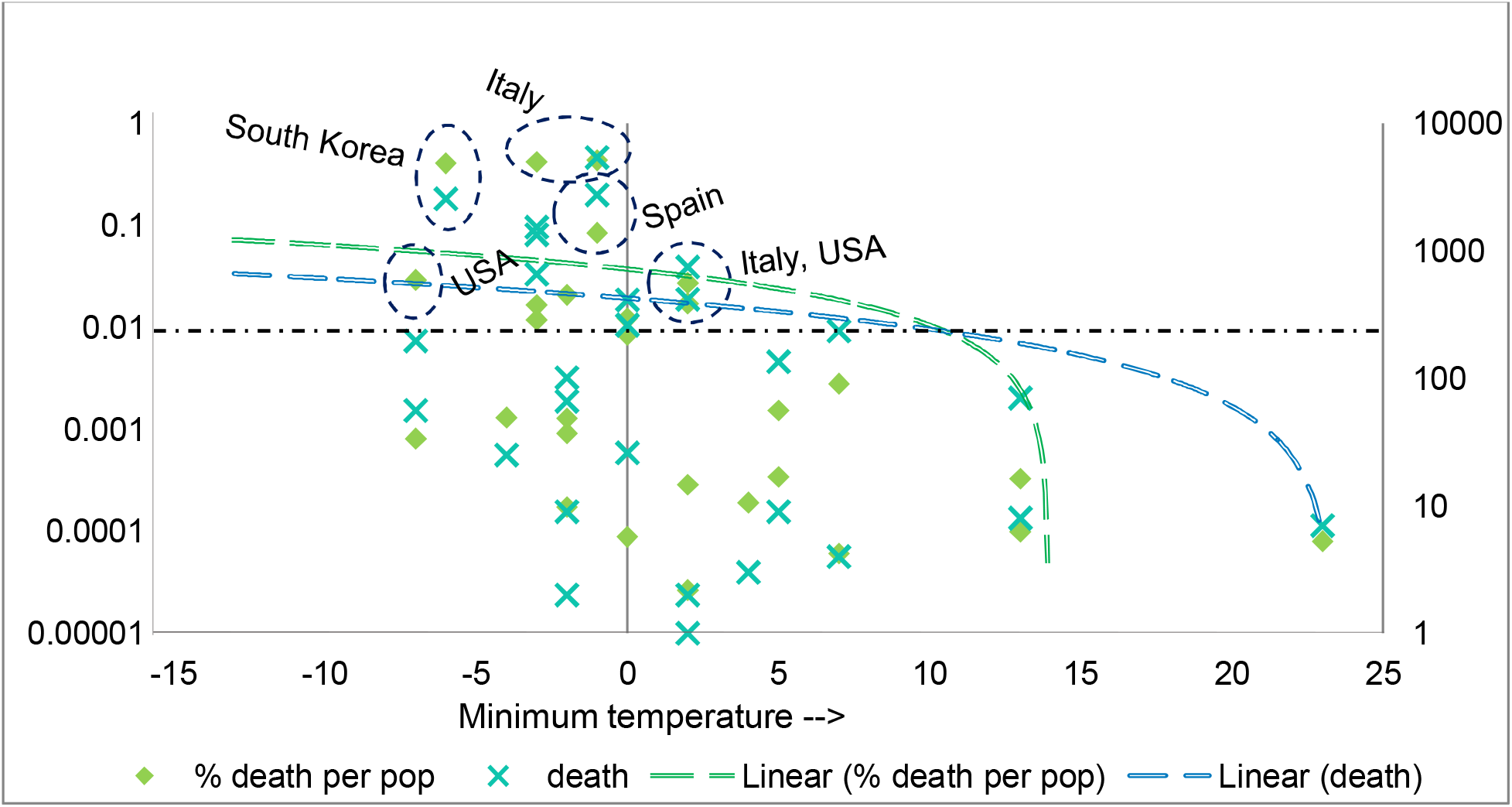
Death with respect to minimum temperature.

On contrary, for highest temperature of a place, a range of the temperatures has shown higher risk of death due to CoVid19. The countries with high death rate had highest day temperature in between 17 to 25 degree centigrade as per figure 2.

**Figure 2:**
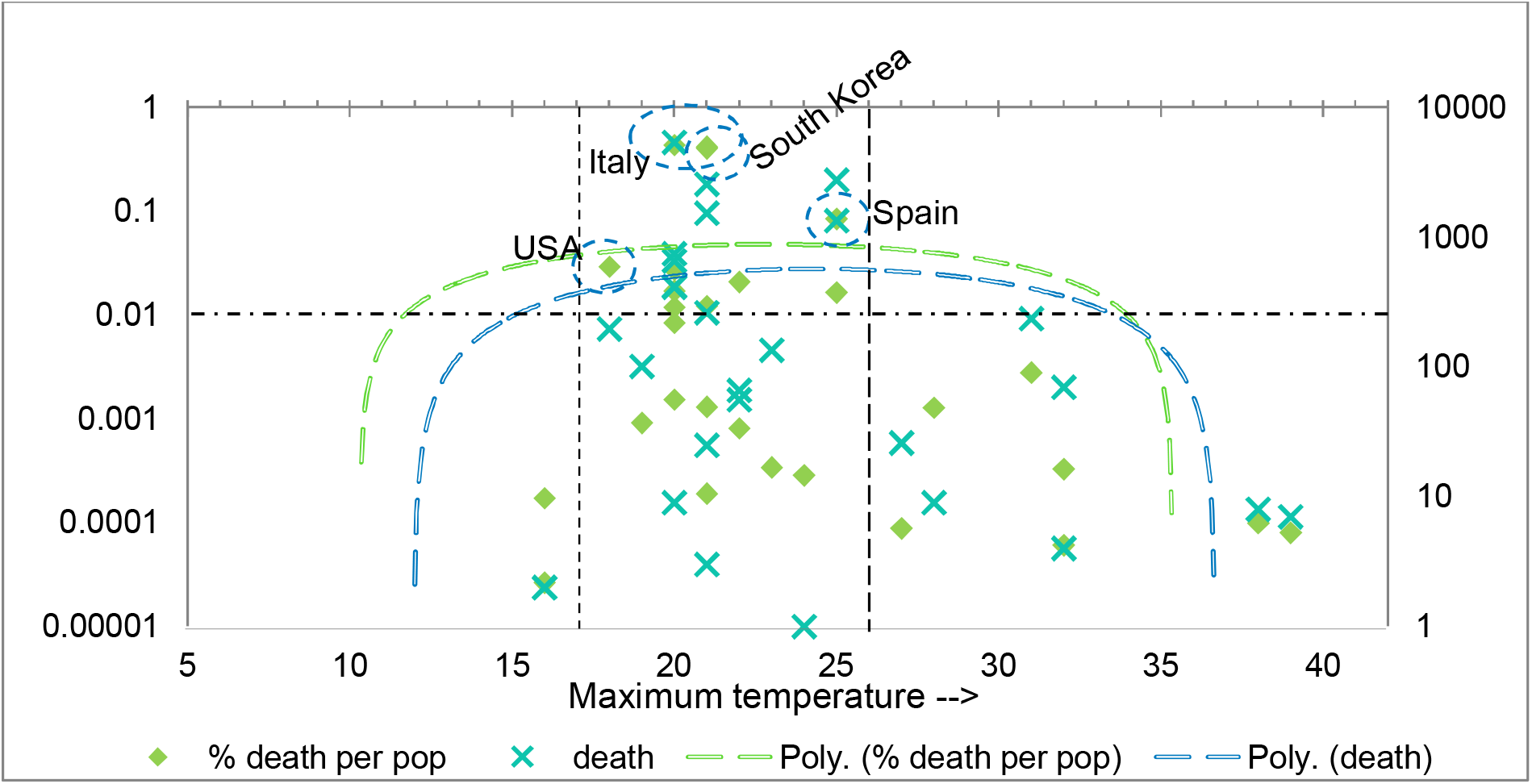
Death with respect to maximum temperature.

### Effectiveness of lockdown

We have given a try to understand the effect of lockdown. How this lockdown is impacting the community transmission is an important study. The entire world is depending on this policy in absence of vaccines. A detailed study of this factor will help us to predict the required lockdown duration for India. We have taken the number of new registered cases of different countries at the starting of lockdown and then after completion of each week till 9^th^ April 2020. We have considered the total change per week to reduce the noise of a temporary change in rate. A sudden change in the number for one or two days without any consistency does not portray any significant change of the overall situation of a country.

The result is presented as graph in figure 3. Only Norway and Italy have entered into the 5^th^ week of lockdown and both countries have shown a drop in new cases after 4^th^ week. But the steadiness is which is yet to be known, and it is highly required to conclude in a positive note. Italy has completed 4 days of 5^th^ week on 9^th^ April 2020. The new cases registered in this duration are 14314, which is a little lower rate than the rate per day of 4^th^ week of lockdown. Austria and Australia have shown notable reduction after 3 weeks of lockdown. After 4^th^ day of the 4^th^ week, the average per day new cases is still very low in Austria. Australia has just completed 1 day of next week as on 10^th^ April. Though per day rate is lower than last week but it is higher for new cases registered in each of the last 2 days. Though no conclusion can be drawn about prevention of infection through lockdown, a prospective evidence of slowdown in the rate of spread of CoVid19 pandemic is available. For a decisive interpretation the impact of lockdown needs to be observed for few more weeks.

**Figure 3:**
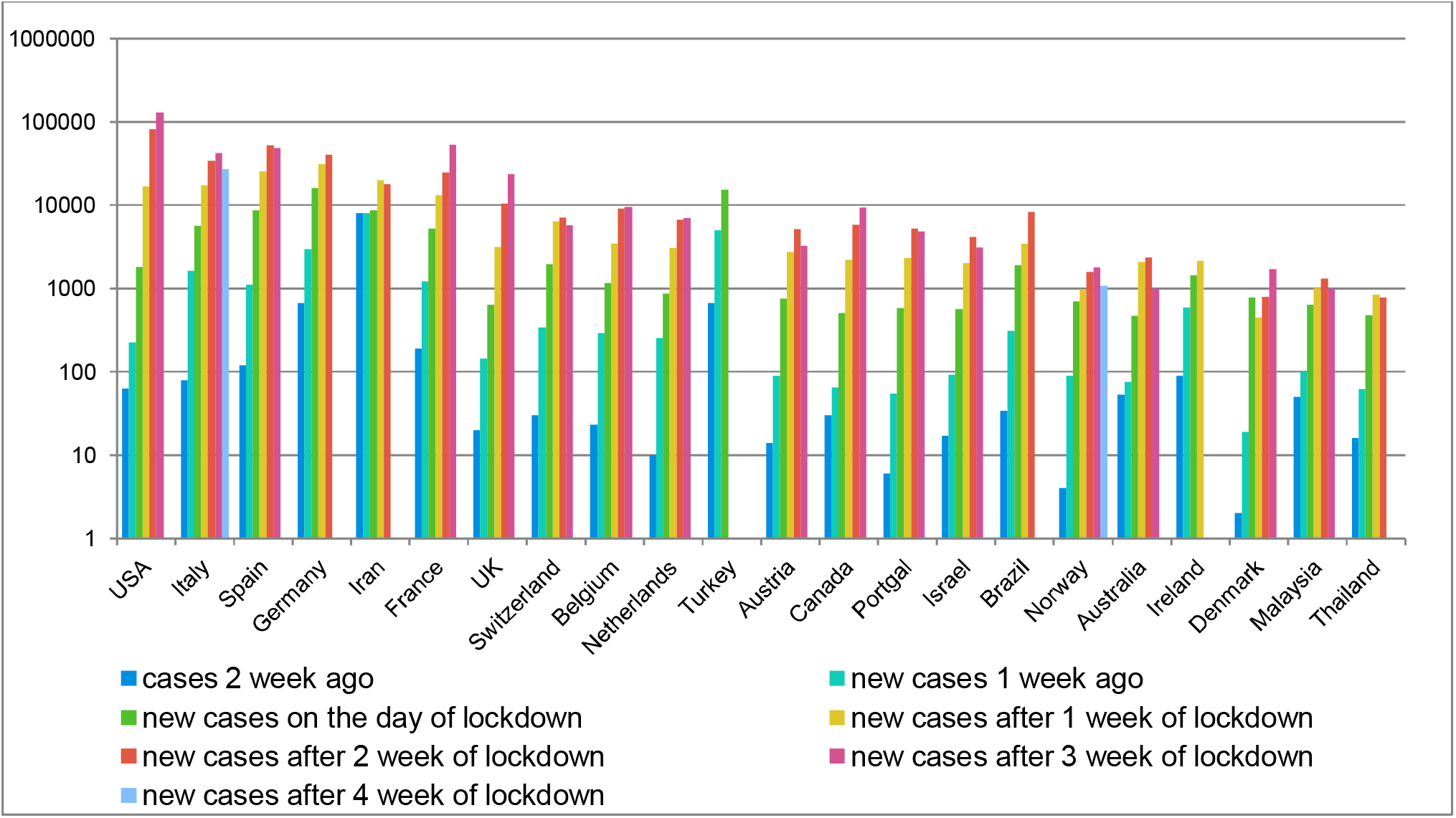
Impact of lockdown.

### Life expectancy and CoVid19

Life expectancy has no direct impact on infection growth and death due to that. But life expectancy is connected with different facts as discussed in introduction. Along with those, depression, low average education and unemployment also have an impact on the life span of a community. To have a measure of all such hidden factors, we have checked the relation between life expectancy and death rate. Here we have taken death per million to compare the change in it with respect to life expectancy. The graph is shown in figure 4. The surprising observation is that, the high death rate is mostly associated with high life expectancy. Inspection of causation is required to understand the actual impact of this finding. This is beyond the scope of our present study and can be considered during advance analysis.

**Figure 4:**
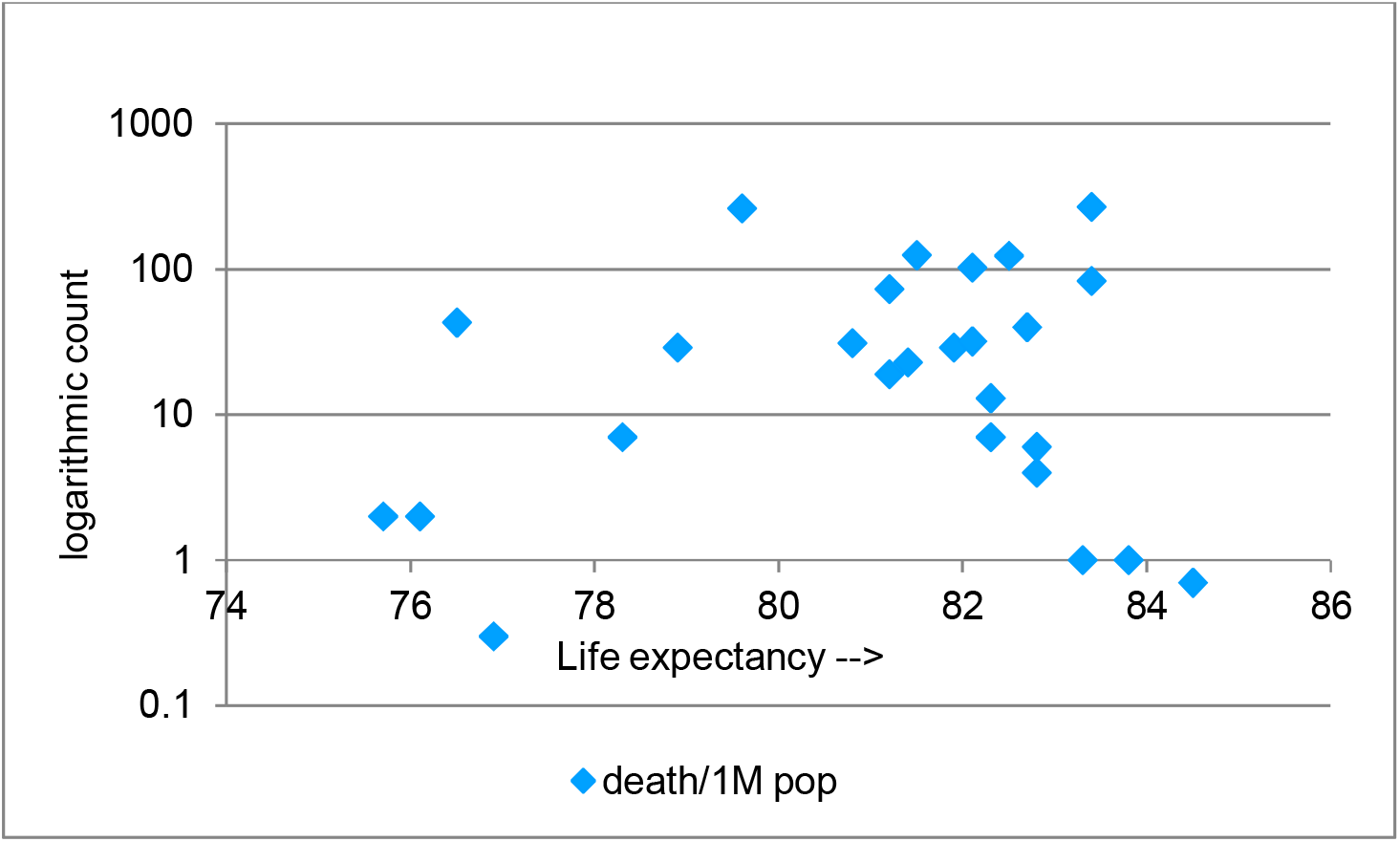
Death rate with respect to life expectancy.

### BCG Vaccination and lungs diseases vs CoVid19

Not all the affected countries are equally prone to CoVid19 as per the report of WHO. Though our preceding analyses have failed to find any strong relationship between the discussed parameters and this pandemic, the presence of some factors influencing the rate of spread of this infection and death rate is obvious. A literature [17] showed some hope in its study of relation between CoVid19 and Bacillus Calmette–Guérin (BGC) vaccination. We have repeated that study to find the relation with present scenario. The work was reported in the middle of March when many countries were not as affected as of today. The scenario in USA has changed dramatically in the last 3 weeks; India also has shown significant increase in the number of infected cases in the last 15 days. Hence, the repetition of the analysis is necessary before concluding any decision.

Our study has shown that vaccination has good impact in most of the cases but there are many exceptions too. The comparison is presented in the figure 5. France, Iran, Ireland, Portugal and Sweden have significant death toll even after having good history of vaccination. On the contrary, Australia and Canada have quite low death rate without vaccination program. Hence the hypothesis of any direct relation between BCG vaccination and CoVid19 has been rejected. A further, in depth analysis including more details of vaccination program, coverage of the population, specially in the below poverty level population, is required to find the exact relation. A low level dependency of death rate on BCG vaccination is visible in the graph which is supported by statistical analysis of correlation resulting in ρ = 0.382 and p-value = 0.054.

**Figure 5:**
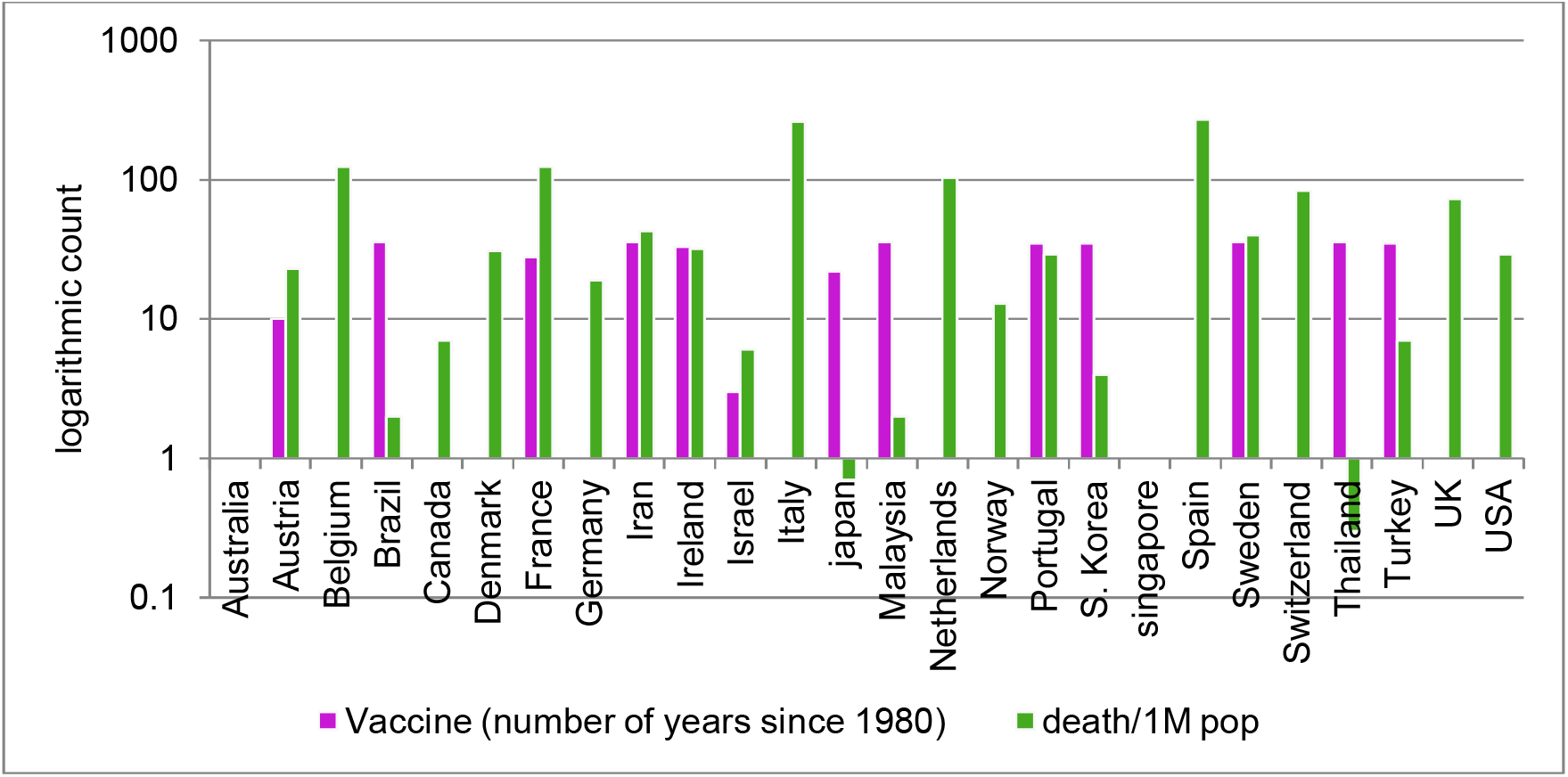
impact of BCG vaccine on death rate.

CoVid19 is a Severe Acute Respiratory Syndrome (SARS) disease. It attacks human lungs, creates trouble in breathing and gradually becomes deadly to claim lives. To understand its relation with other lungs disease, a study has been conducted. Top 10 diseases of each country are examined to see the burden of other lung diseases. After preliminary scrutiny, we decided to consider lung cancer (LC), Chronic obstructive pulmonary disease (COPD) and Lower respiratory infect (LRI). Data cleaning and preprocessing was done before analysis. The rank of these diseases which are among the top 10 and primarily responsible for death in a country has been considered for further analysis. If a disease is not listed in a country’s top 10 diseases, a numerical value 0 has been considered. The correlation and p-value of these 3 diseases with death rate are ρ = 0.535, p-value = 0.06. The p-value depicts a higher tendency towards null hypothesis though the correlation is average. The relation LC alone is better with death rate. With a ρ = 0.477, p-value = 0.013 LC demands a command over death rate due to CoVid19. This finding does not offer any way of prevention of the pandemic but gives an idea about the risk of a country.

## Discussion

None of the above discussed parameters has significant effect CoVid19 except lungs cancer. Though few have shown impact in increase in number of cases (or deaths) in some cities, nothing convincing has been found. It seems like the disease gets transmitted with almost equal potential in different atmosphere. Impact of lockdown is also not considerably good to get any definite suggestion. Hence the question remains unsolved. How India is going to response to this pandemic?

Negative minimum temperature, a specific range of maximum temperature, lack of BCG vaccination and tendency of other lungs diseases have shown some positive impact in increasing the number of CoVid19 cases and death. We have combined all these four parameters to see their combined effect on death rate. Before statistical analysis, we have done preprocessing of each parameter to create four distinguished features.

The temperature data are analogous in nature having no significant impact in case of a minute change. Using the already acquired knowledge from the previous analysis, we have classified the temperature into two different classes. In case of minimum temperature, values equal to or below 0°C are considered as one class which has strong negative impact on death and hence represented by 10. Rest of the temperature values which have less impact or no impact on death are considered as 1. Similarly, maximum temperatures between 17 to 25°C are converted to 10 and rest are to 1. Disease scores are divided into 3 classes - 10 to 6, 5 to 1 and 0. Score greater than 5, represents disease rank within top 5. For these ranks a disease is represented by 10. Scores from 5 to 0 means the disease is ranked between 6 to 10 and rank below 5 is presented by 5. When a disease is not listed in top 10 diseases of the corresponding country, it has been represented by 0. This data preprocessing steps are important for better insight. Vaccination is represented by number of the years the program is continued in a country since 1980 as mentioned before.

After creating a clean dataset with these four features, statistical analysis was done to check if any useful correlation is present. This analysis offers a prominent correlation, ρ = 0.634 with high acceptance, p = 0.024.

The features are plotted in figure 7 along with increasing death rate. In the figure, Singapore, Australia and France have shown exception in vaccination impact. The probable reason can be the temperature. Both Singapore and Australia are hot countries. Australia is not prone to any lungs disease and Singapore has low tendency of LC. On the other hand, France has low temperature and high impact of LC in country’s death rate. Hence, a significant impact of these parameters can be assumed, and this needs further research for definite conclusion.

**Figure 6:**
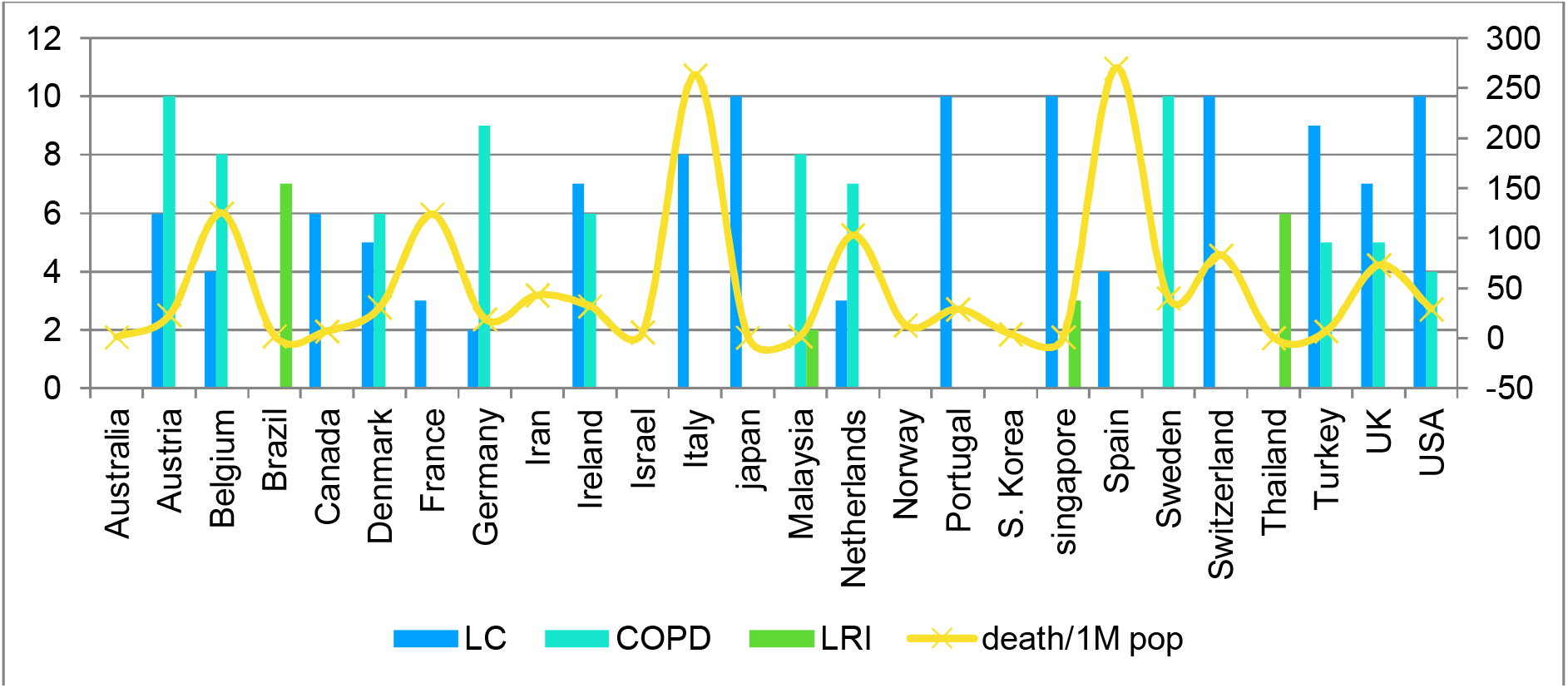
Lungs diseases vs death rate.

**Figure 7:**
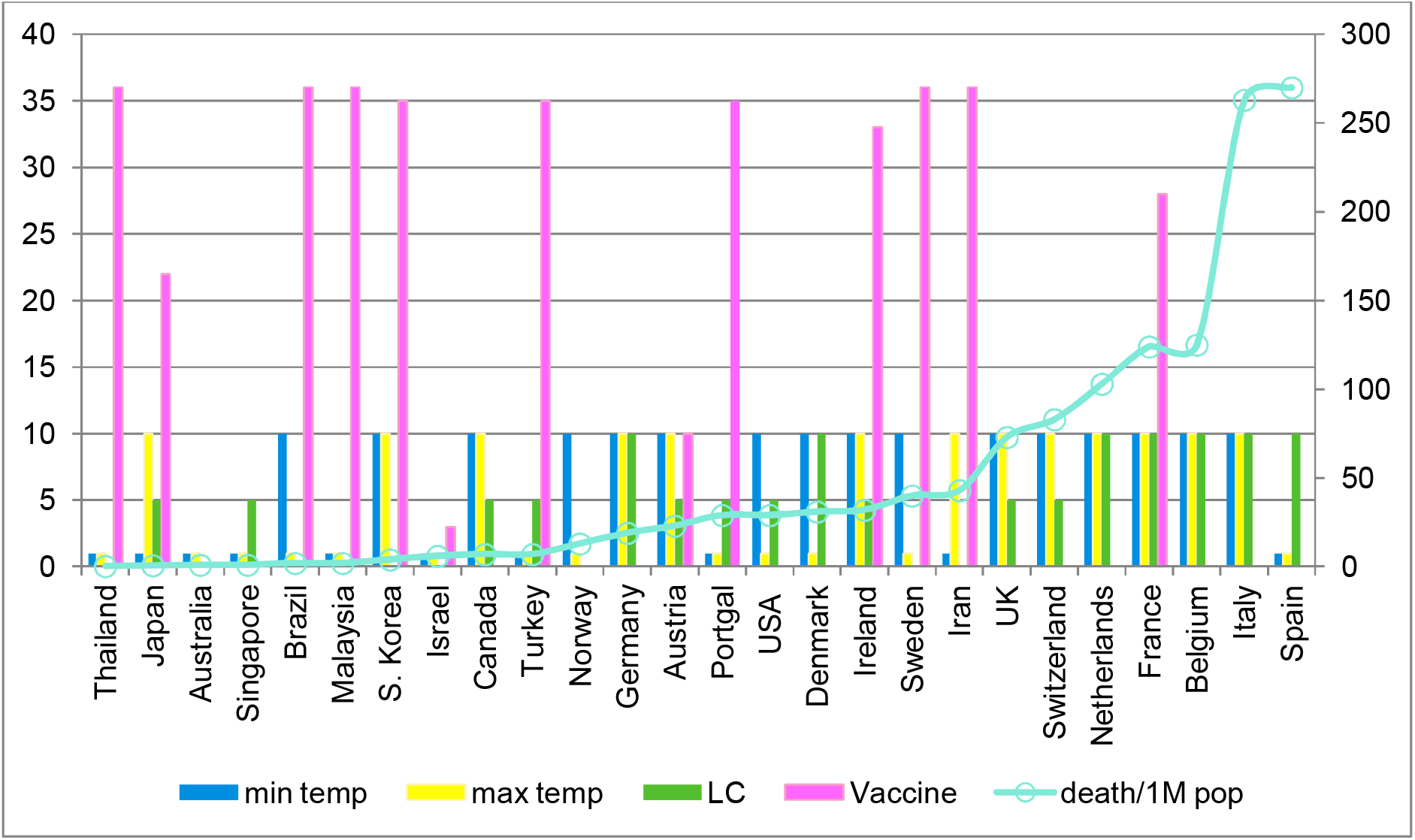
selected features and death rate.

In April, temperature remains significantly high in India (non-hill zones). In most of the areas, specially in the cities which are reporting high rate of cases and death, the lowest temperature remains higher than 15°C and maximum temperature goes well above 30°C. India also have different vaccination program for years and BCG vaccination is done for almost 90% of the population. The negative factor is that the lung diseases are very common in India. COPD (rank 2), LRI (rank 5) and tuberculosis (rank 6) are major causes of death here, though LC is not that common like other countries which are badly affected by CoVid19 pandemic.

This information and data dependent statistical analysis is not self-sufficient to understand the nature of coronavirus. Along with this geographic, demographic and meteorological analysis, information from other branches of science like virology, biotechnology must be considered. An important finding about such pandemic is their sudden disappearance after few months. It happened every time in the world’s history. Not considering the expected life of SARS-Cov2 with respect to environmental conditions and continuous mutation will be impractical. The faster the virus will self-limit itself, the earlier the rate of infection will go down.

## Conclusion

India has imposed quarantine through locked down for 21 days starting from 23^rd^ of March 2020. In last 15 days the number of cases as well as death has increased significantly. The lockdown do restrict the community transmission but the rate is increasing may be because of detection of already infected cases. Fast identification of old cases is required to access the effect of isolation on infection transmission rate. The mortality and morbidity ratio may be affected by the immune system of a population and will vary with different life style factors starting from food habits, common diseases, vaccination programs etc.

In the high altitude areas (mainly the Himalayan region) with low temperature throughout the year, the risk is higher as per our analysis. February, March are not tourism season of these Himalayan region of India because of chilling cold, road blockage due to snowfall and for academic session ending with examinations. Probably due to little tourist flow, these regions are still not reporting cases of CoVid19. Once the severity of pandemic will fall and the country will start resuming its normal life, the free movement of tourists can be a threat for those hilly areas and it could ignite a reappearance of the disease in cities too through the returned tourists. A proper protection plan and restricted movement for reasonably longer time after the pandemic is required to keep the citizens safe.

To summarize the risk of India with more accuracy, we have to consider the socioeconomic condition too. A good vaccination program, history of having seasonal endemics ensures better immunity against similar diseases. SARS-CoV-2 is a new virus and hence the old antibodies cannot completely prevent it. But whether any antibody has any significant impact on its growth or not is under research. Cyclicity is ubiquitous for acute infectious diseases [23]. Each disease is unique on its own. The SARS diseases occupy the span of two months, March and April, of Indian epidemic calendar. The reason is that the seasonal variation has significant impact on transmission of infectious diseases. This is known as seasonal forcing. If the vaccine of tuberculosis is resistive for CoVid19, then the already present antibodies in the blood will reduce the severity of the disease and death rate in India.

## Data Availability

Attached as file

## Authors’ contribution

The first author, Soumi Ray, Ph.D. in Image analysis from the Indian Institute of Technology Roorkee, has done the data analysis, explored useful insights, prepared the manuscript and concluded the work. The second author, Mitu Roy, B.Tech in Computer Science from Haldia Institute of Technology, conceived the content and retrieved the data to help the first author.

## Funding

None.

## Conflict of interest

Both the authors declare to have no conflict of interest.

## Acknowledgment

None.

## Notes

### Competing Interest Statement

The authors have declared no competing interest.

### Funding Statement

NO fund

